# Toward Predicting Peripheral Artery Disease Treatment Outcomes Using Non-Clinical Data

**DOI:** 10.1101/2024.08.05.24311525

**Authors:** Ali Al Ramini, Farahnaz Fallahtafti, Mohammad Ali TakallouIraklis, Iraklis Pipinos, Sara Myers, Fadi Alsaleem

## Abstract

Peripheral Artery Disease (PAD) significantly impairs quality of life and presents varying degrees of severity that correctly identifying would help choose the proper treatment approach and enable personalized treatment approaches. However, the challenge is that there is no single agreed-on measure to quantify the severity of a patient with PAD. This led to a trial-and-error approach to deciding the course of treatment for a given patient with PAD. This study uses non-clinical data, such as biomechanical data and advanced machine-learning techniques, to detect PAD severity levels and enhance treatment selection to overcome this challenge. Our findings in this paper lay the groundwork for a more data-driven, patient-centric approach to PAD management, optimizing treatment strategies for better patient outcomes.

## Introduction

This paper presents the use of non-clinical data to characterize severity levels and support decision-making when treating chronic diseases such as peripheral artery disease (PAD). PAD is an atherosclerotic syndrome that leads to occlusion of the arteries supplying the legs. PAD affects approximately 8 million people in the US, producing a considerable public health burden.^1–3^ The standard therapies for PAD include pharmacotherapy,^4^ supervised exercise therapy^5^, wearing assistive devices^6,7^ endovascular revascularization (angioplasty/stenting), and open revascularization (bypass operations).^8^ Knowing which treatment will benefit each patient most is very challenging. Clinical evidence shows that treatment outcomes vary widely across patients with PAD^4^, and the factors contributing to the success or failure of treatments are poorly understood.^1,9^ For example, while assistive ankle foot orthoses (AFO) show promise in enhancing walking distances, the research on its consistent use and patient perceptions is limited.^10^ Conventional pharmacological treatments do not address existing blockages and muscle myopathy experienced by individuals with PAD and only minimally improve walking distances.^11,12^ Revascularization lacks clear superiority between bypass surgery and endovascular revascularization, and surgical interventions, in general, have high risks to patients and varying patency rates.^13-17^ All these limitations highlight the need for evidence-based guidelines for treating a patient with PAD.^18^

An additional challenge in PAD treatment is that there are no standard agreed-upon measures to confirm improvement after treatment. Existing outcome measures vary, and the most common measures are not inclusive by only assessing blood flow improvements (ankle-brachial indices) or very subjective, such as patient questionnaires or self-reported improvement or worsening of symptoms. These limitations, when combined with PAD’s complex nature, have limited physician’s ability to understand which course of treatment for a given patient with PAD would lead to the best outcomes.^3^

A predictive model can significantly enhance clinicians’ ability to make informed decisions, tailor interventions, and optimize post-treatment care of patients with PAD, thus improving care quality and reducing ineffective or high-risk interventions.^19^ Toward this goal, this paper seeks to harness the power of machine learning and comprehensive gait analysis to provide a proof of concept for a data-driven approach to PAD treatment and management. Our approach leverages the data we have collected for patients with PAD over the last 20 years. The comprehensive dataset includes clinical data and gait biomechanics measurements for healthy subjects and patients with PAD before and after treatments. Our recent findings demonstrated that a machine learning approach with gait biomechanics data could accurately classify individuals as having or not having PAD (Figure 1.a).^20,21^ Building on this finding, our current study will use comprehensive biomechanics gait data to establish reliable measures for the PAD severity level (Figure 1.b), which can then be used to develop models predicting a specific treatment outcome (Figure 1.c). These models are the first step toward building an intelligent expert system to support individualized treatment decisions for patients with PAD (Figure 1.d).

**Figure 1:**
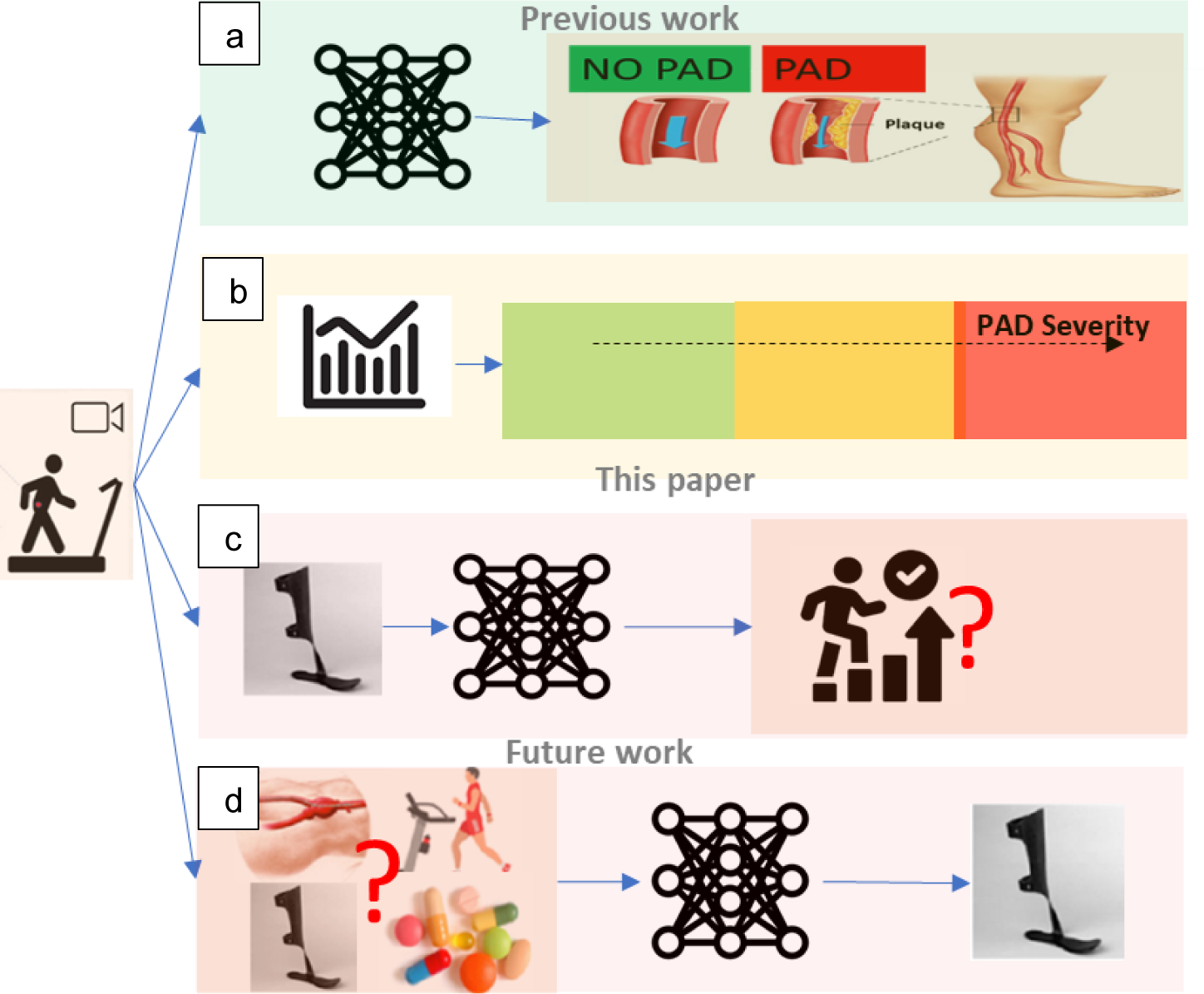
Our Progress and future work toward PAD treatment prediction. a) Our previous work showed that biomechanics data could be used to classify individuals as having or not having PAD.^20,21^ b) This paper aims to establish a measure of PAD severity, which will be the engine to c) train models to predict treatment outcomes of patients with PAD and then in the future to d) predict the optimal treatment for patients with PAD.

## Results

Figure 2. a presents a comprehensive distribution of all features in our study for healthy controlled and patients with PAD before different treatments (AFO and Surgery). The features are represented after the post-application of the Yeo-Johnson transformation. It is evident from the figure that kinematics features (ankle, hip, and knee) failed to differentiate between healthy individuals and those with PAD. While qualitative questionnaire evaluations, such as SF-36 and WIQ, effectively distinguished between healthy controls and patients with PAD, they failed to discern PAD severity, particularly when differentiating between AFO and surgery patients. The severity (supported by clinical measures such as the ABI test) is ranked based on the type of intervention performed on a patient with PAD: Healthy - AFO - Surgery. In other words, the fact that a patient with PAD has undergone open surgery means a vascular physician determined his PAD severity level to be higher than the patient who had just AFO. The GRF features distinguished all three categories and presented an ordered patient distribution. This observation highlights the potential of GRF features as a potent instrument for quantifying PAD severity.

**Figure 2:**
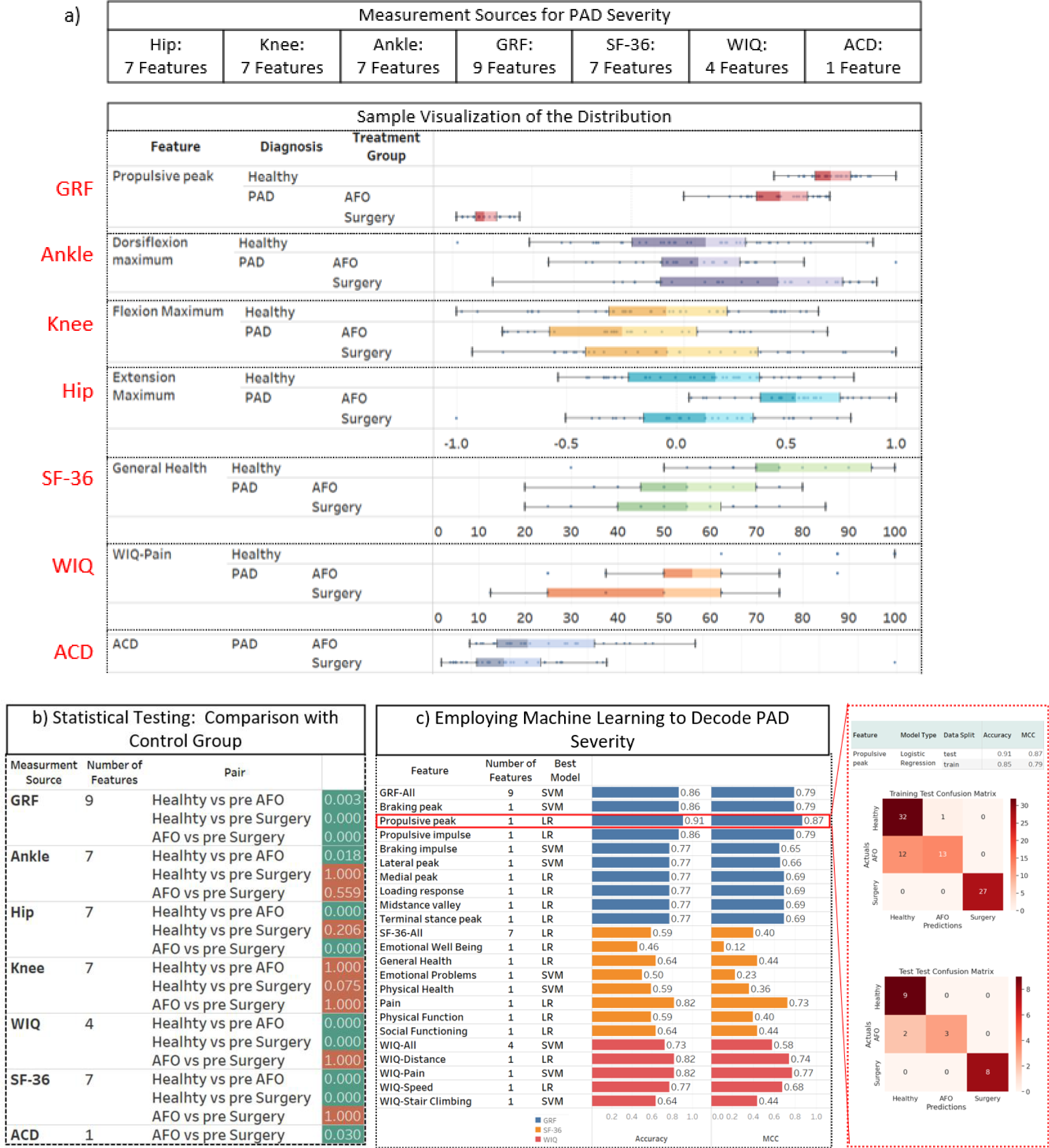
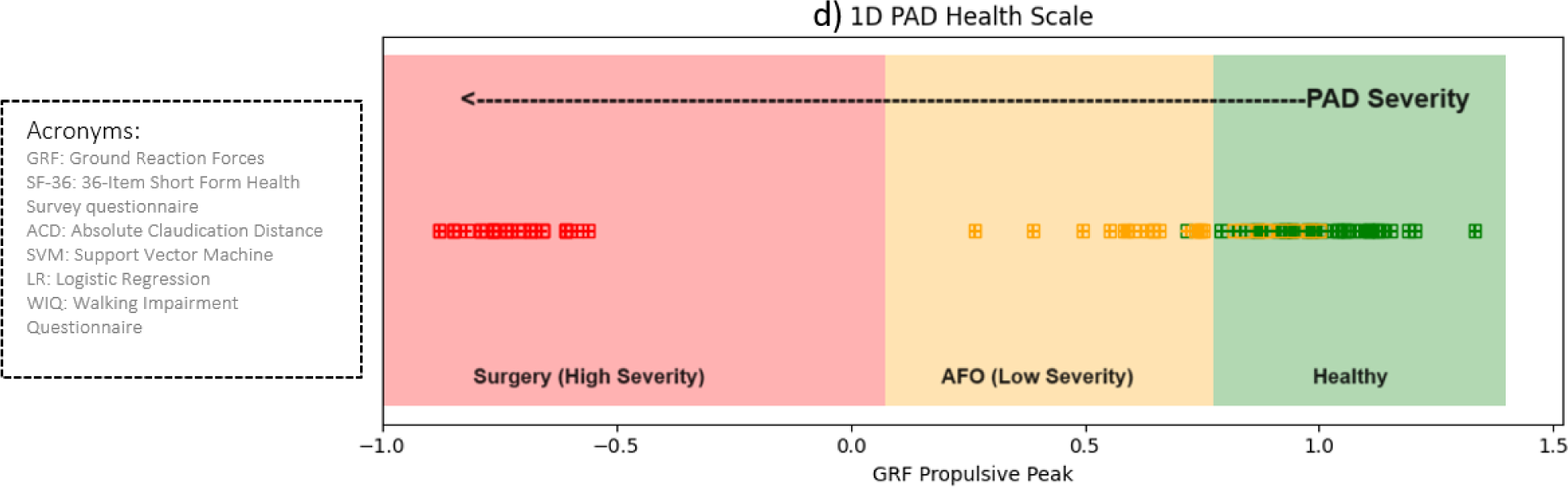
GRF emerges as a strong indicator of PAD severity. a) Pre-intervention distribution of all features categorized based on diagnosis and treatment groups (Healthy, AFO, and Surgery). b) Results of the Mann-Whitney U test comparing the Healthy, AFO, and Surgery groups across various gait features. The figure showcases each measurement source’s Median Bonferroni P Value, with lower values indicating higher statistical significance. c) Comparative performance of machine learning models in feature-level PAD severity quantification across all features as independent inputs. d) Stratification of PAD Severity Based on GRF Propulsive Peak Values: A continuous scale derived from Logistic Regression model predictions, delineating three distinct regions - Healthy (-1 to -0.8), AFO (Less Severe PAD, -0.8 to 0.25), and Surgery (More Severe PAD, 0.25 to 1). This scale correlates GRF Propulsive Peak values and PAD severity, facilitating swift and precise clinical assessments.

Figure 2.b presents the outcomes of the Mann-Whitney U test aiming to recognize statistically significant differences among the Healthy, AFO, and Surgery groups. The results confirm the observations (Figure 2.a) that GRF features emerge as most able to distinguish across groups, exhibiting the most significant statistical differences between the three groups in the pre-intervention phase. In contrast, hip, ankle, and knee kinematic features do not consistently capture the disparities between these groups. Furthermore, the WIQ questionnaire features effectively differentiate between the Healthy group and patients with PAD groups, but not between the AFO and Surgery groups.

Next, the results of using machine learning modes to decode PAD severity using the different features are shown in (Figure 2.c). The GRF, using logistic regression with a single feature, “Propulsive peak,” achieved an accuracy^22^ of 0.909, a balanced accuracy^22^ of 0.867, and a Matthews Correlation Coefficient (MCC)^22^ of 0.868. The GRF’s high performance across these metrics, particularly the MCC that accounts for unbalanced data, highlights its effectiveness in accurately quantifying PAD severity. Additionally, the consistent results between the model’s training and testing performance suggest that the GRF situation effectively captures the essential patterns in the data without being overly fitted to the training set or missing the situation’s complexity. This confirms the robustness of GRF features for reliable severity assessment in PAD.

Figure 2.d aims to provide a more granular understanding of PAD severity by leveraging the GRF Propulsive Peak, the best-performing feature identified in the previous analysis, by stratifying a PAD scale. The data analysis yielded a discernible stratification of PAD severity based on GRF Propulsive Peak values (Figure 2.d) derived from the logistic regression model predictions. The resulting figure defines three distinct regions:

- Surgery Region (More Severe PAD): Ranging from -1 to 0.075.
- AFO Region (Less Severe PAD): Extending from 0.075 to 0.775.
- Healthy Region: Above 0.775 on the GRF Propulsive Peak scale.

This stratification visually represents how GRF Propulsive Peak values correlate with varying degrees of PAD severity. In summary, the results in Figure 2 show the importance of GRF as a robust and reliable data source for PAD severity quantification, outperforming other commonly used metrics and questionnaires such as SF-36 and WIQ.

Next, interest is shifted to understanding the patients with PAD’s response to interventions using effect size analysis^23^. This analysis quantitatively represents the strength of the relationship between variables using Cohen’s d measure. Cohen’s guidelines suggest that a d value of 0.2 indicates a ‘small’ effect size, 0.5 a ‘medium’ effect size, and 0.8 a ‘large’ effect size. In our context, with a control group and two patient groups (Surgery and AFO), both pre and post-intervention, the effect size aids in quantifying the magnitude of change due to intervention in each patient group and offers a comparative measure against the control. For instance, comparisons such as Healthy vs. pre-surgery provide insights into the deviation of the surgery group from the control before the intervention. At the same time, Healthy vs. post-surgery reveals the deviation post-intervention.

Figure 3.a presents the effect sizes, quantified using Cohen’s d, for one GRF measure and four WIQ measures as an example relative to the healthy control group. Figure 3.a shows the magnitude of this effect size based on GRF Propulsive Peak, confirming the severity level differences between patients with PAD who had surgery versus patients completing an AFO intervention. More importantly, the effect size decreases post-surgery, suggesting a shift towards the healthy group’s values. In contrast, the AFO group shows minimal change in the GRF values post-intervention. Both behaviors are somewhat expected, extending the confirmation that GRF can also be used to capture PAD status after intervention. However, the WIQ fails to capture the anticipated severity levels among patients with PAD following AFO and surgery. In addition, their effect size indices project potentially over-optimistic results, with post-surgical and post-AFO cohorts yielding equivalent Cohen’s d magnitudes.

**Figure 3:**
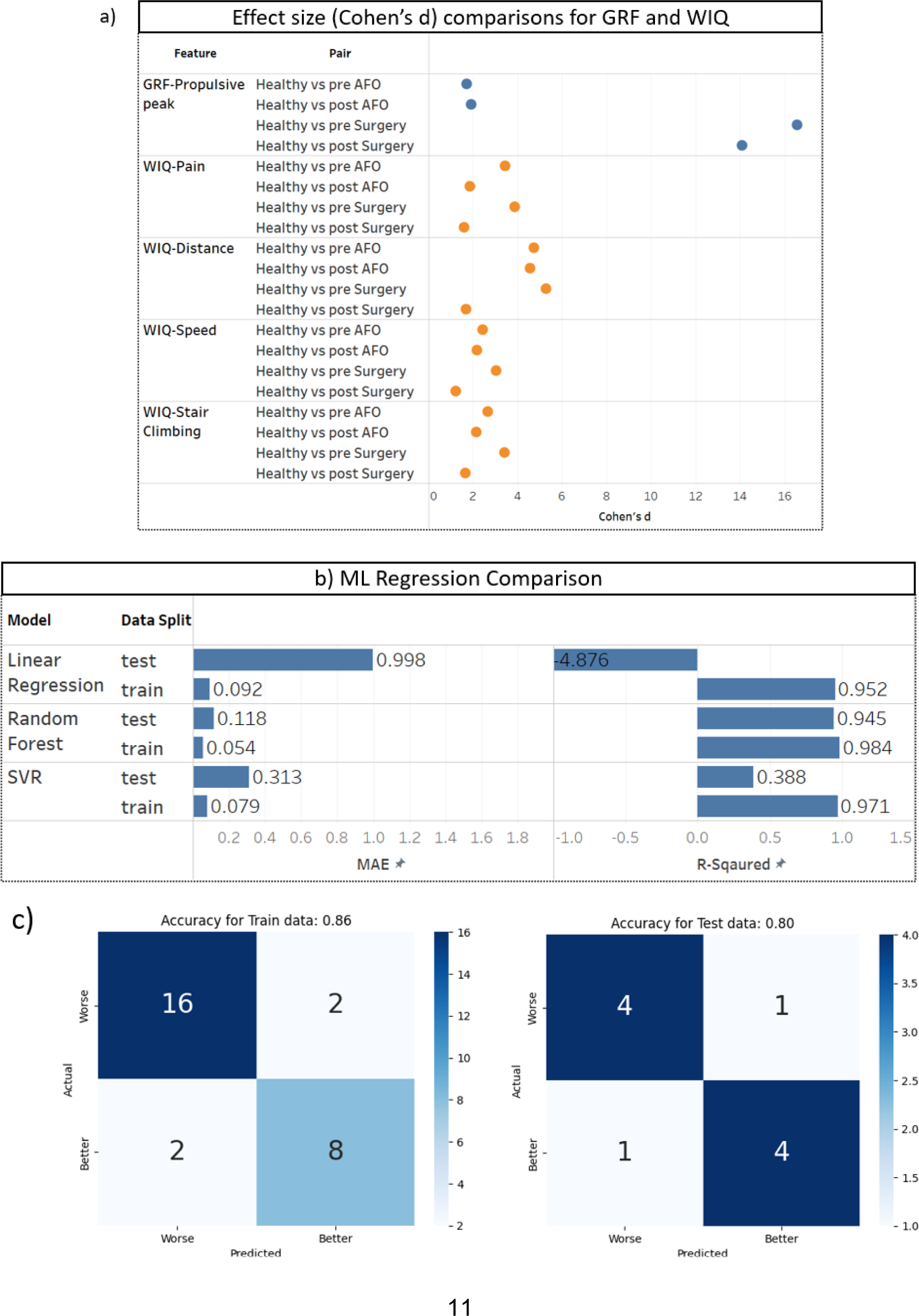
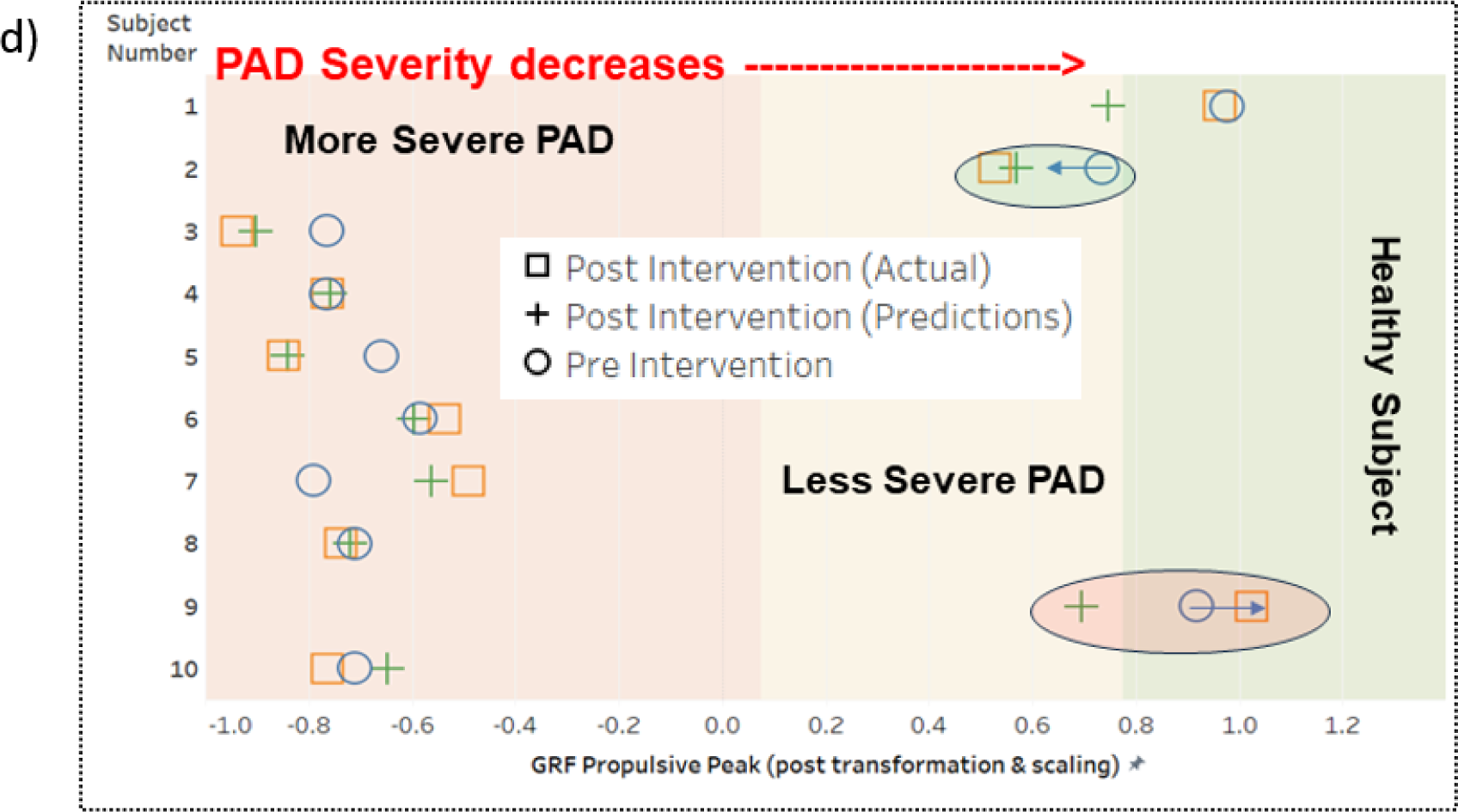
Machine learning models are developed to predict the outcome of a certain treatment. a) Effect size (Cohen’s d) comparisons for GRF, WIQ, and SF-36 scores between healthy individuals and patients with PAD, both pre and post-intervention. Higher Cohen’s d values indicate larger differences between the groups, suggesting more pronounced effects of interventions or more distinct group characteristics. Blue dots represent comparisons involving the ‘pre’ status, and orange dots signify the ‘post’ status. b) Performance metrics comparison of machine learning models including Linear Regression, Random Forest, and SVR. Metrics shown are MAE, R-Squared, and adjusted R-Squared, assessing model accuracy and predictive capability with both Correlation and PCA feature selection methods on training and test datasets. c) Confusion matrices for treatment outcome predictions (Worse vs. Better) using the Random Forest model on training and test datasets, showing the counts of correct and incorrect predictions. d) Individual Predictions of GRF Propulsive Peak for All Subjects Post-Intervention. The graph plots the actual vs. predicted GRF Propulsive Peak values, illustrating prediction accuracy. The spectrum of ‘More Severe PAD’ to ‘Less Severe PAD’ visualizes the range of severity based on GRF values, with a clear demarcation showing patients transitioning towards a ‘Healthy’ status. Notably, the distribution of cases in ‘More Severe’ and ‘Less Severe’ PAD is not purely based on the number of surgeries or AFO interventions but on the observed severity metrics in the GRF Propulsive Peak values post-treatment.

Next, the most profound part of this paper is that machine learning models were trained using pre-intervention data, including gait kinematics and kinetics measurements, WIQ scores to predict post-intervention effectiveness. Post-intervention effectiveness is captured by improvement in GRF Propulsive Peak. Figure 3b results provide a comprehensive overview of the performance metrics for various machine-learning models (Linear Regression, Random Forest, and SVM). Figure 3.c shows that the Random Forest model consistently outperforms the other models in Mean Absolute Error (MAE) and R-squared. Furthermore, we added a binary representation for the model outcomes by assessing if the predicted values can specify whether a patient’s situation improved based on the predicted and actual data results. The confusion matrices for both training and test datasets, as depicted in Figure 3.c, provide a clear visual representation of the model’s predictions against the actual outcomes. For the training data, the model achieves an accuracy of 86%, while for the test data, the accuracy stands at 82%. These high accuracy scores indicate the model’s ability to assess treatment outcomes’ direction. This indicates that machine-learning applications could be practical in predicting treatment outcomes for PAD.

The individual predictions of GRF Propulsive Peak post-intervention for each subject, as shown in Figure 3.d, offer a granular view of the model’s performance. Each subject’s data point is plotted against their pre-intervention GRF Propulsive Peak, with distinct markers indicating the actual post-intervention values and the model’s predictions. A consistent alignment between the actual and predicted values is evident across most subjects. This individual-level analysis complements the accuracy metrics, reinforcing the model’s ability to predict treatment outcomes. The colored bars visually represent the model’s confidence in its predictions, with the proximity of the actual and predicted markers indicating the precision of the model’s forecasts.

## Discussion

The inability of simple tests to differentiate between healthy controls and patients with PAD emphasizes the need for more robust metrics in estimating a patient with PAD severity level. While qualitative assessments like SF-36 and WIQ have their merits, their inability to discern between AFO and Surgery patients raises questions about their efficacy. Remarkably, the GRF Propulsive Peak feature stands out, with the ability to differentiate across all categories and an ordered distribution among patients with PAD that suggests their potential as a foundation of characterizing PAD severity. The GRF Propulsive Peak introduces a straightforward 1D scale for measuring PAD severity. This 1D scale is precious for its practical applications outside clinical settings, offering a simple and effective tool for monitoring PAD severity. The 1D scale is a beacon for future innovations that aim to make PAD management more efficient and patient-centric.

GRF Propulsive peak values using the effect size test show a better correlation with post-treatment interventions. On the other hand, qualitative assessments such as WIQ scores might present an overly optimistic view of the patient’s condition. This asserts the importance of integrating GRF measurement with subjective patient-reported outcomes to understand PAD severity and its response to interventions.

The Random forest model accurately predicts the post-intervention GRF propulsive peak. Using such a model, clinicians can gain valuable insights into the potential success of a treatment before its actual implementation. This predictive capability can revolutionize patient care, allowing for more personalized treatment plans and potentially reducing the number of ineffective interventions. Moreover, it is a foundational step towards a more data-driven approach in PAD treatment, where decisions are informed by predictive analytics rather than merely relying on traditional methods. As we continue to refine and validate this model, it paves the way for more comprehensive studies that can further unravel the complexities of PAD and optimize treatment strategies for better patient outcomes.

The ability to predict individual treatment outcomes with high accuracy, as demonstrated by the Random Forest model, holds significant implications for clinical practice. Clinicians can effectively tailor their treatment approaches and post-treatment care by estimating the post-intervention GRF Propulsive Peak. The model’s performance, both in terms of overall accuracy and individual predictions, suggests that it can be a valuable tool in decision-making. As we delve deeper into personalized medicine, such predictive capabilities become increasingly crucial. The alignment between the model’s predictions and the actual outcomes underscores the potential of integrating machine learning into PAD treatment strategies. This integration enhances the precision of treatment planning and facilitates more informed patient-clinician discussions, fostering a collaborative approach to care.

While providing valuable insights, our study has certain limitations. Firstly, the dataset employed is relatively constrained, encompassing only 97 subjects, of which 42 are healthy controls, and 65 are patients diagnosed with PAD. Consequently, our analytical and machine learning outcomes necessitate validation through a broader dataset with a greater patient count and severity variability. Secondly, the result encapsulated in the GRF Propulsive peak scale offers room for refinement. The current scale predominantly categorizes patients with PAD into two severity brackets: less severe (AFO patients) and more severe (Surgery patients). A more nuanced representation could be achieved by incorporating a broader spectrum of PAD severity gradations.

Future research can add more patients with PAD data from other treatments to the model to improve generalization and provide more options for predicting treatment outcomes. Moreover, real-time GRF data could provide a more nuanced understanding of PAD severity and post-treatment effect.

## Methods

### Data Sources and Description

Biomechanics^24^ data for this study were sourced from research approved by the Institutional Review Boards at the University of Nebraska Medical Center and the Nebraska-Western Iowa Veteran Affairs Medical Center, involving 65 individuals with PAD and 42 healthy controls. Of the patients with PAD, 9 were treated with AFO, and 35 underwent surgical treatments. Some AFO-treated patients’ data were collected multiple times pre-intervention. Therefore, we have 30 records for AFO-treated patients before intervention. Not all patient data were collected after the intervention; 30 surgery patients’ data were collected after the surgery, and 8 AFO-treated patients’ data were collected after three months of the treatment. In this study, we considered patients treated with AFOs to have less severe PAD than those treated with surgery.

In addition to biomechanical data, qualitative and quantitative assessments were conducted to understand the PAD condition better. The WIQ was administered to gauge the self-perceived walking ability of patients with PAD^25-27^. This questionnaire evaluates various aspects of walking, including pain, distance, speed, and stair climbing, providing insights into patients’ daily challenges with PAD. SF-36 was also employed to assess the participant’s health status and quality of life^12,28^. This comprehensive questionnaire evaluates physical and mental health domains, offering a broader perspective on the impact of PAD on patients’ daily lives.

Furthermore, the Absolute Claudication Distance ^26^test was conducted. This quantitative assessment measures the distance a patient can walk before being compelled to stop due to claudication pain, providing a direct metric of the severity of PAD symptoms. Collectively, these assessments, combined with the biomechanical data, aimed to offer a multi-dimensional perspective on PAD’s impact on patients. Furthermore, this multi-dimensional dataset offers an opportunity to compare various measurement sources to identify the most precise metric for assessing PAD severity.

For this analysis, multiple measurement sources were considered:

1. Ankle, Hip, and Knee Kinematics: Each anatomical region was characterized by seven distinct features, capturing the nuances of movement and biomechanical alterations.^29^
2. Ground Reaction Forces (GRF): Nine features were extracted to understand the forces exerted during walking, providing insights into gait alterations.^24^
3. Walking Impairment Questionnaire (WIQ): Four features were derived from this self-administered questionnaire, offering a patient-centric perspective on walking ability.^26^
4. Medical Outcomes Study Short Form 36 (SF-36): Seven features were considered from this tool, gauging the participants’ health status and quality of life.
5. Absolute Claudication Distance (ACD): This singular feature was exclusively available for patients with PAD, measuring the distance they could walk before the claudication pain.

### Data Transformation

In the preliminary stages of our analysis, we identified the need to standardize and transform our biomechanical data to ensure comparability across various biomechanics features and conditions. We employed the Yeo-Johnson transformation^30^, a method optimized for varying data distributions, including zero and negative values. Initially, this transformation was determined and performed based on the pre-intervention dataset. Subsequently, the same transformation parameters derived from the pre-intervention data were applied to both the post-intervention and healthy datasets, ensuring consistency across all conditions. After the transformation, we implemented feature scaling to standardize the data distribution further, which is crucial for the performance of certain machine learning models and the validity of statistical tests that assume data uniformity. This standardization facilitates more accurate analyses and helps in achieving reliable results. This scaling was configured using the transformed pre-intervention data, ensuring that all biomechanical features ranged between -1 and 1. The same scaling parameters were then applied to the post-intervention and healthy data. This systematic data transformation and scaling approach enhanced data clarity, separation, and comparability across all conditions and features.

### Statistical Comparison with Control Group Using the Mann Whitney U-test

We employ the non-parametric Mann-Whitney U-test^31^ to distinguish the differences in measurements between the three independent groups - Healthy, AFO, and Surgery. The Healthy group comprised individuals without PAD, serving as the control, while the AFO (low severity) and surgery (high severity) groups represented patients with PAD varying in degrees of disease severity.

We chose The Mann-Whitney U-test due to its robustness in comparing non-normally distributed data, which is often the case in medical research. In addition, many features in our data have shown non-normality. We present the results from this test as the Median Bonferroni P-value for each measurement source, which adjusts the median of the observed p-values to account for multiple comparisons, reducing the chances of type I errors in our statistical analysis.^32^ It is worth noting that the ACD data was only available for patients with PAD. Thus, comparisons involving the healthy group did not apply to this measure.

This statistical approach aims to identify the measurement source that best differentiates between the three groups. Ideally, the most informative measurement source would be the one that exhibits significant differences across all three pairwise comparisons: Healthy vs. AFO, Healthy vs. Surgery, and AFO vs. Surgery. For instance, if the GRF features consistently show significant disparities among these three comparisons, it would suggest that GRF features are the most potent identifiers of PAD severity.

### Machine learning approach to evaluate features’ ability to quantify PAD severity

In this section, we take a more granular approach to PAD severity classification by focusing on individual features within each data source—GRF, WIQ, and SF-36. Each feature is modeled and tested separately. This allows us to isolate the predictive power of each feature, providing a more subtle understanding of its role in PAD severity classification.

The data is initially segmented into individual features and then subjected to a machine learning pipeline involving data splitting, data preprocessing, hyperparameter tuning, model training, and performance evaluation. We continue to employ Logistic Regression^33,34^, Decision Tree^35^, and Support Vector Machine (SVM)^36^ models for this analysis. The goal is to identify the most informative individual features for PAD severity quantification, offering insights into the potential for streamlined, feature-centric diagnostic approaches. This feature-level assessment aims to refine our interpretation of which aspects of GRF, WIQ, and SF-36 most relate to PAD severity. It could thus be a focal point in future diagnostic and severity assessment tools.

**Figure 4:**
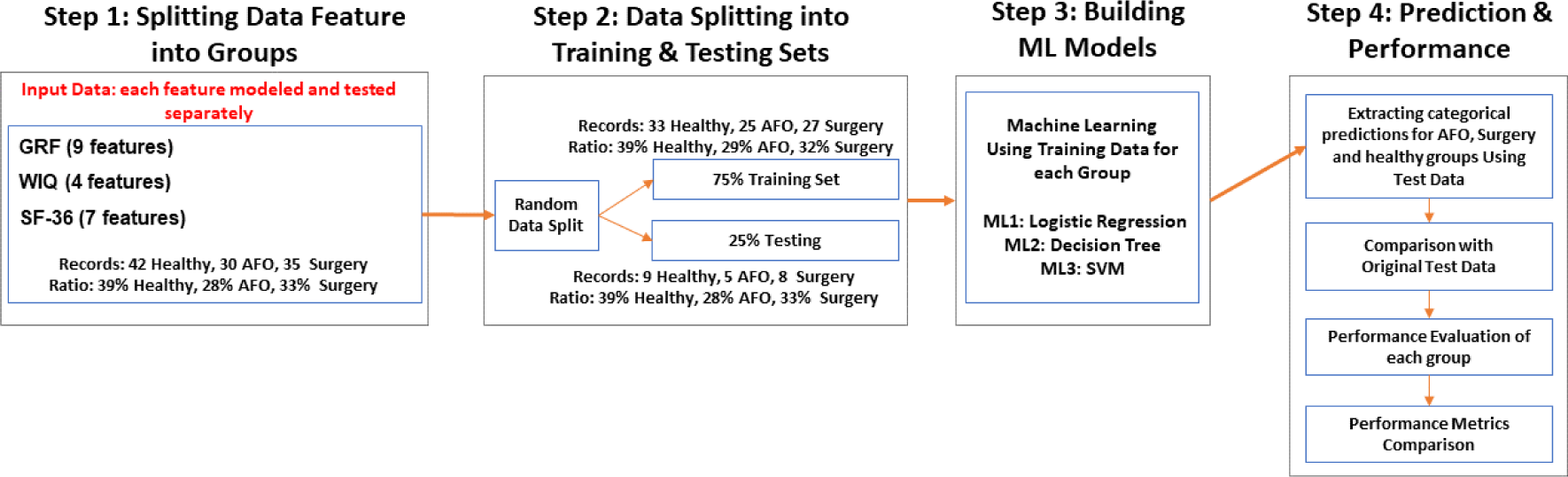
Flowchart overview of feature-level assessment for PAD severity quantification. The chart compares individual features using logistic regression, decision tree, and SVM models.

### Stratification of PAD Severity Using Optimal Features

This section aims to provide a more granular understanding of PAD severity by leveraging the best-performing feature identified in the previous analysis. This 1D scale could be a valuable reference for future engineering applications to monitor PAD severity and progression outside of the clinical environment, mainly because this feature is derived from GRF that can be physically measured in a lab setting. We stratify a 1D PAD scale based on the Logistic Regression model built using the GRF Propulsive Peak feature described in Figure 2.c. We synthesized a hypothetical dataset to elucidate the relationship between “GRF Propulsive Peak” values and PAD severity. This hypothetical dataset spanned ‘GRF Propulsive Peak’ values from -1 to 1.4, incremented by 0.025. Leveraging our optimal Logistic Regression model, previously identified as the most potent single feature model for this task, we processed this dataset to generate PAD severity predictions. We categorized these predictions into our multiclass classification groups: Healthy, AFO, and Surgery. The objective was to establish a continuous scale that could intuitively represent PAD severity based on GRF Propulsive Peak values.

### Quantifying Intervention Impact Using Effect Size

Effect size^23^ is a quantitative representation of the strength of the relationship between variables. Unlike statistical tests that merely confirm the existence of an effect or relationship, the effect size elucidates its magnitude, independent of sample size. In our context, with a control group and two patient groups (Surgery and AFO), both pre and post-intervention, the effect size aids in quantifying the magnitude of change due to intervention in each patient group and offers a comparative measure against the control.

We employ Cohen’s d, a measure calculated as the difference between two means divided by the pooled standard deviation to quantify the differences between our groups. For instance, comparisons such as Healthy vs. pre-surgery provide insights into the deviation of the surgery group from the control before the intervention. At the same time, Healthy vs. post-surgery reveals the deviation post-intervention. Analogously, comparisons for the AFO group, Healthy vs. pre-AFO and Healthy vs. post-AFO provide similar insights. Cohen’s guidelines suggest that a d value of 0.2 indicates a ‘small’ effect size, 0.5 a ‘medium’ effect size, and 0.8 a ‘large’ effect size. The sign of Cohen’s d further provides directional information: a positive sign indicates the post-intervention group has higher scores than the pre-intervention or control group. In contrast, a negative sign indicates the opposite. This approach aims to provide a nuanced, quantitative understanding of PAD severity and its response to interventions.

### Machine Learning Approach Estimating Post-Treatment Outcomes

The section utilizes pre-intervention data, including gait kinematics and kinetics measurements, WIQ scores, and ACD as predictive features. The initial step involves gathering pre-intervention data from a cohort of 38 patients diagnosed with PAD. This data is sourced from various measurements, including gait kinematics covering ankle, hip, and knee features and gait kinetics focusing on ground reaction forces (GRF), WIQ, and ACD. Additionally, patients are categorized based on their treatment group: AFO for those with low severity and surgery for those with high severity. Out of the total, 30 patients underwent surgery due to their high severity, while eight were treated with AFO due to their lower severity.

The data is split into training and testing sets. The training set comprises 28 patients, with six from the AFO group and 22 from the surgery group. The remaining ten patients, consisting of 3 AFO and seven surgery patients, are reserved for the test set. This division allows the model to be trained on a diverse data set and subsequently validated on unseen data to gauge its predictive accuracy.

Before feeding the data into machine learning models, it undergoes a feature selection process to enhance its predictive capability. The method employs Principal Component Analysis (PCA)^37^ for dimensionality reduction. PCA transforms the original features into a set of linearly uncorrelated variables, capturing the most significant patterns in the data while reducing its complexity (90% of the total variance is maintained). We applied this feature selection method at the training data source level to ensure that every data source contributed to the study while avoiding data leakage between the train and test sets.

**Figure 5:**
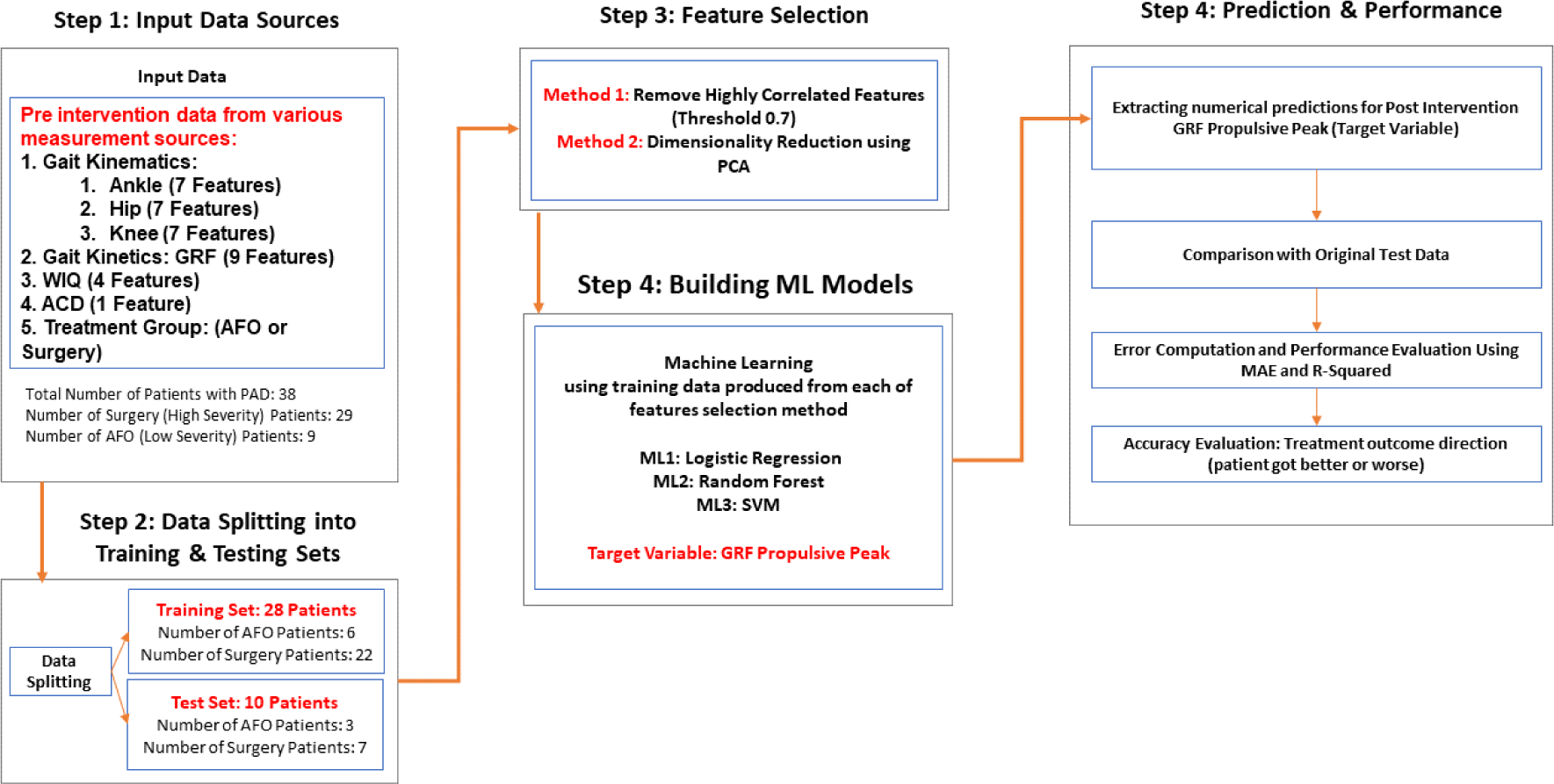
Machine learning flowchart. The chart depicts the machine learning approach for predicting PAD treatment outcomes using pre-intervention data and evaluating model performance.

Three machine learning models were trained on the features of Linear Regression, Support Vector Regression (SVR), and Random Forest^38^. These models were chosen for their versatility and capability to handle complex nonlinear relationships in the data. The trained models were then used to predict the GRF Propulsive Peak post-intervention, which, as established in previous chapters, serves as a measure of PAD severity and a reference for treatment outcome.

Three machine learning models are trained with optimized features: Logistic Regression, Random Forest, and Support Vector Machine (SVM). Each model is trained using the data produced from both feature selection methods. Post-training, the models predict the post-intervention GRF Propulsive Peak (established in the previous chapter, which is the most distinctive measure for PAD severity), serving as the target variable. These predictions are then compared with the original test data. The model’s performance is evaluated using MAE for average prediction error magnitude and R-squared for the proportion of variance explained by the model. Additionally, the accuracy of the treatment outcome direction, indicating whether a patient’s condition improves or worsens, is assessed.

## Data Availability

Data is available per request and after approval

